## Supplementary figures and images for "Prevalence of anti-SARS-CoV-2 antibodies in Poznań, Poland, after the first wave of the COVID-19 pandemic"

### Supplemental Figure 1

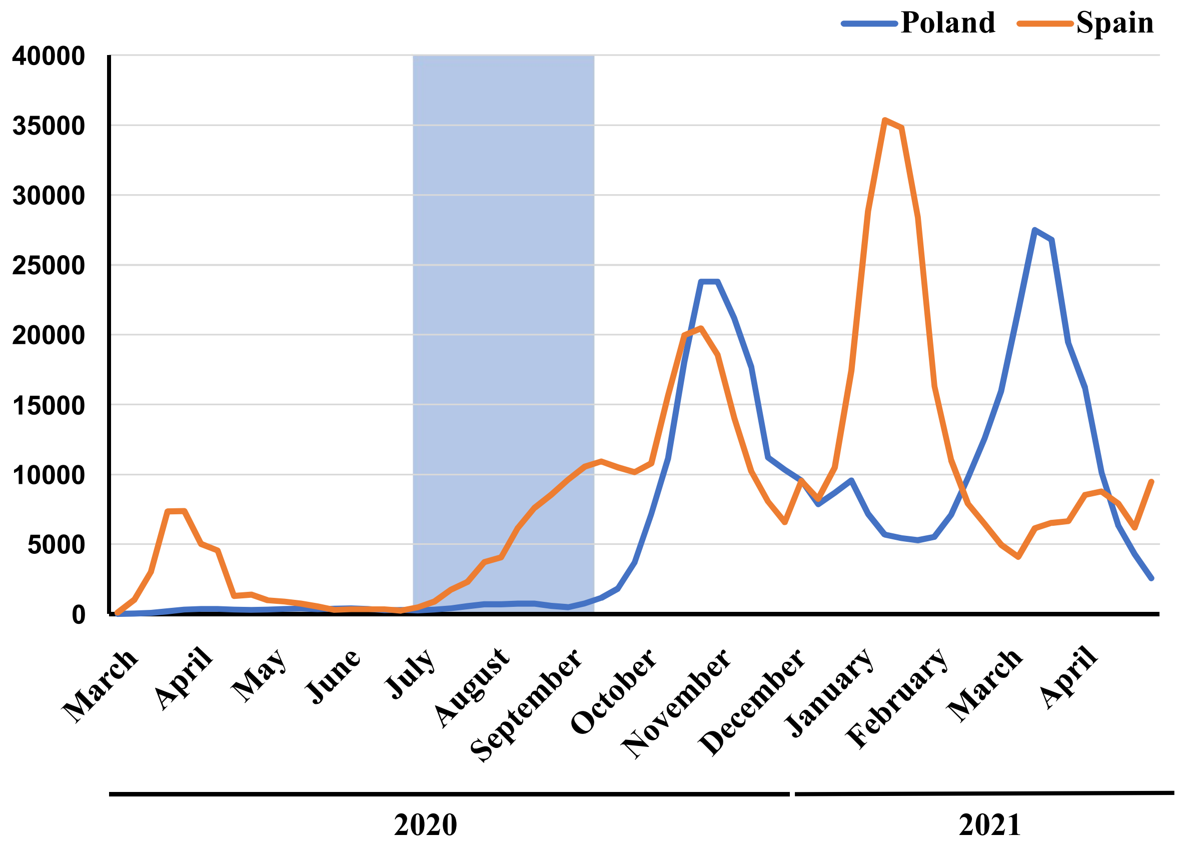
